# Use of serology immunoassays for predicting SARS-CoV-2 infection: a serology-based diagnostic algorithm

**DOI:** 10.1101/2021.10.23.21265429

**Authors:** Alejandro Lazo-Langner, Benjamin Chin-Yee, Jaryd Tong, Lori Lowes, Benjamin D. Hedley, Michael Silverman, Johan Delport, Vipin Bhayana, Michael Knauer, Ian Chin-Yee

**Affiliations:** Division of Hematology, Department of Medicine, Western University, London, Ontario, Canada; Department of Epidemiology and Biostatistics, Western University, London, Ontario, Canada; Department of Pathology and Laboratory Medicine, Western University, and London Health Sciences Centre, London, Ontario, Canada; Division of Infectious Diseases, Department of Medicine, Western University, London, Ontario, Canada

**Keywords:** serology, SARS-CoV2, COVID-19, diagnosis, prediction, algorithm

## Abstract

**Background:** Detection of viral RNA by nucleic acid amplification testing (NAAT) remains the gold standard for diagnosis of SARS-CoV-2 infection but is limited by high cost and other factors. Whether serology-based assays can be effectively incorporated into a diagnostic algorithm remains to be determined. Herein we describe the development of a serology-based testing algorithm for SARS-CoV-2 infection.

**Patients and Methods:** Between July 2020 and February 2021, we included symptomatic unvaccinated patients evaluated in the Emergency Department of our institution for suspected SARS-CoV-2. All patients had testing by real-time Reverse Transcription Polymerase Chain Reaction. The performance characteristics of five commercial enzymatic serology assays testing for different antibody isotypes were evaluated in a derivation cohort and the assay with the best performance was further tested on a validation cohort. Optimal cut-off points were determined using receiver operating characteristic (ROC) curves and further tested using logistic regression.

**Results:** The derivation and validations cohorts included 72 and 319 patients, respectively. Based on its initial performance, the Elecsys Anti-SARS-CoV-2 assay (Roche Diagnostics) was further tested in the validation cohort. Using ROC curve analysis, we estimated the diagnostic performance for different cut-off points assuming a prevalence of positive tests of 5%. At any given cut-off point the NPV was over 97%.

**Discussion:** This study suggests that an initial diagnostic strategy using the Elecsys Anti-SARS-CoV-2 serology test in symptomatic unvaccinated patients could help to rule out an acute SARS-CoV2 infection and potentially lead to appropriately tailored infection control measures or rational guidance for further testing with a potential cost reduction and increased availability.

## 1. Introduction

Effective diagnosis of SARS-CoV-2 infection remains a cornerstone of clinical and public health responses to the current pandemic. Since the initial outbreak of the COVID-19 pandemic in December 2019, significant advances have been made in the diagnosis, management and prevention of SARS-CoV-2 infection (1-3). Detection of viral RNA by nucleic acid amplification testing (NAAT) remains the gold standard for diagnosis of SARS-CoV-2 infection. However, NAAT-based testing is complicated by high cost, technical complexities and scalability which can all lead to long turn-around-time, often exceeding 24 hours. Complex technical requirements, including technical expertise, instruments availability, as well as accessibility to large scale collection centres limit its implementation, particularly in resource-constrained settings, thus impacting clinical management and infection control measures. Considering the limitations, there has been a proliferation of alternative testing methods for SARS-CoV-2 infection, primarily in the form of serology-based assays, which offer advantages of lower cost and more rapid turn-around-time relative to NAAT (4, 5). While serology-based testing has been applied in some healthcare settings, more widespread adoption remains limited by lack of clarity of performance relative to gold standard NAAT-based testing, and failure to incorporate within diagnostic algorithms for patients with suspected SARS-CoV-2 infection. We previously evaluated the performance of five SARS-CoV-2 serology assays in samples from patients tested for SARS-CoV-2 infection by NAAT, demonstrating high sensitivity and specificity of several serology-based assays(6). Whether specific cut-off values for serology-based assays can accurately predict negative NAAT testing and be effectively incorporated into a diagnostic algorithm remains to be determined. Herein we describe the development of a serology-based testing algorithm for SARS-CoV-2 infection based on a cohort of patients presenting with symptoms of acute infection who underwent both serologic- and NAAT-based testing. We describe an approach using up-front serology testing to predict negative NAAT result and effectively rule out SARS-CoV-2 infection in symptomatic patients.

## 2. Materials and methods

### 2.1 Study Design and population

This study was conducted at a tertiary care centre in Ontario, Canada, after approval by the institutional Research Ethics Board. The derivation phase consisted of a retrospective cohort of patients assessed between March and April 2020. The validation phase consisted of a prospective cohort including patients assessed between July 2020 and February 2021. We included patients evaluated in the Emergency Department (ED) for suspected symptomatic SARS-CoV-2 infection and who underwent NAAT testing, for whom clinical information was available and had available stored serum or plasma samples collected for other purposes around the time of NAAT testing. We collected demographic and clinical data including age, sex, comorbidities, time to symptom onset, and laboratory data. We also documented duration of symptoms at time of NAAT testing for SARS-CoV-2 infection.

### 2.2. SARS-CoV-2 testing

Serology and NAAT testing were performed in independent laboratories in a blinded fashion. All patients included had testing by real-time Reverse Transcription Polymerase Chain Reaction (rRT-PCR), using a research-use only E-gene / EAV assay (Cat No. 40-0776-96, TIB Molbiol Syntheselabor GmbH, Berlin, Germany) and RNA Virus Master (Cat No. 06754155001, Roche Diagnostics International Ltd., Rotkreuz, Switzerland). Extraction and amplification were respectively performed on the Hamilton STAR (Hamilton Company, Reno NV, USA) and Roche LightCycler 480 II instruments according to manufacturer’s instructions. In the derivation cohort serology was tested using five commercially available assays. The EUROIMMUN Anti-SARS-CoV-2 ELISA IgA and ELISA IgG assays were performed on an EUROIMMUN Analyzer-I (EUROIMMUN Medizinische Labordiagnostika AG, Lübeck, Germany). These are semiquantitative assays for the detection of IgG or IgA antibodies against the Domain S1 of the SARS-CoV-2 spike protein and are reported as ratios (Negative <0.8). The DiaSorin’s LIAISON SARS-CoV-2 S1/S2 IgG assay was performed on the Liaison XL instrument (Diasorin S.p.A., Saluggia, Italy). This is a quantitative assay for the detection of IgG antibodies against the S1/S2 antigens and results are reported as arbitrary units per millilitre (Negative <12). The Epitope Diagnostics Novel Coronavirus COVID-19 IgM assay (Epitope Diagnostics Inc., San Diego, CA, USA) is a qualitative assay designed to detect multiple epitopes of the SARS-CoV-2 nucleocapsid protein. This test was manually performed in a Multiskan™ FC Microplate Photometer (ThermoFisher Scientific, Waltham MA, USA) and reported as a ratio based on optical density at 450 nm (Negative ≤ 0.9). The Elecsys Anti-SARS-CoV-2 was performed on an automated Cobas e801 analyzer (Roche Diagnostics International Ltd., Rotkreuz, Switzerland). It is a qualitative electro-chemiluminescence immunoassay (ECLIA) for the detection of total antibodies against the nucleocapsid antigen reported as a cut-off index based on the provided calibrators (Negative <1). Serology-based assays were performed on peripheral blood serum or plasma samples collected at the time of NAAT testing, according to manufacturers instructions. These assays have demonstrated high sensitivity and specificity for detection of SARS-CoV-2 infection in prior studies(6) and have a rapid turn-around-time on high volume automated ELISA analyzers.

### 2.3. Data analysis

In the derivation cohort baseline characteristics were compared for NAAT-positive and negative samples using Pearson Chi-square, Student’s t, or Mann-Whitney tests, as appropriate. Single and multiple variable stepwise logistic regression analyses were conducted to determine the strength of association between each potential predictor and a positive NAAT test. Results for all serology tests were dichotomized using receiver operating characteristic (ROC) curves to determine optimal cut-off points for each test’s reporting units using Youden indices. Logistic regression models were constructed to determine the strength of association of dichotomous serology results with positive NAAT testing and adjusted for the time differential between NAAT and serology testing. Goodness-of-fit was assessed using Hosmer-Lemeshow tests and models were internally validated using non-parametric bootstrapping with 1,000 iterations. Pre-specified subgroup analyses were conducted using regression models for samples with a time differential between NAAT and serology testing of plus or minus 48 hours. For all dichotomous serology cut-off values, we calculated sensitivity, specificity, and negative likelihood ratios. Negative predictive values (NPV) were calculated assuming a prevalence of positive tests of up to 5%.

For the validation cohort, subsequent serology testing was chosen based on tests’ procedural considerations and statistical performance. Logistic regression models were constructed as previously described and were further adjusted for time from symptoms onset to testing. Sensitivity ROC curve analyses were conducted for different cut-off points and for the subgroup of samples with a time differential between testing within 48 hours. All analyses were conducted using SPSS 22.0 software (IBM Corp., Armonk NY, USA) and MedCalc Statistical Software version 19.2.6 (MedCalc Software Ltd, Ostend, Belgium).

## 3. Results

### 3.1. Patient characteristics

The derivation and validations cohorts included 72 and 319 patients, respectively. All patients presented to the ED with symptoms and underwent testing by NAAT. Approximately half of all patients were male, and the mean age was approximately 65 years. Patient characteristics are summarized in **Table 1**.

**Table 1.**
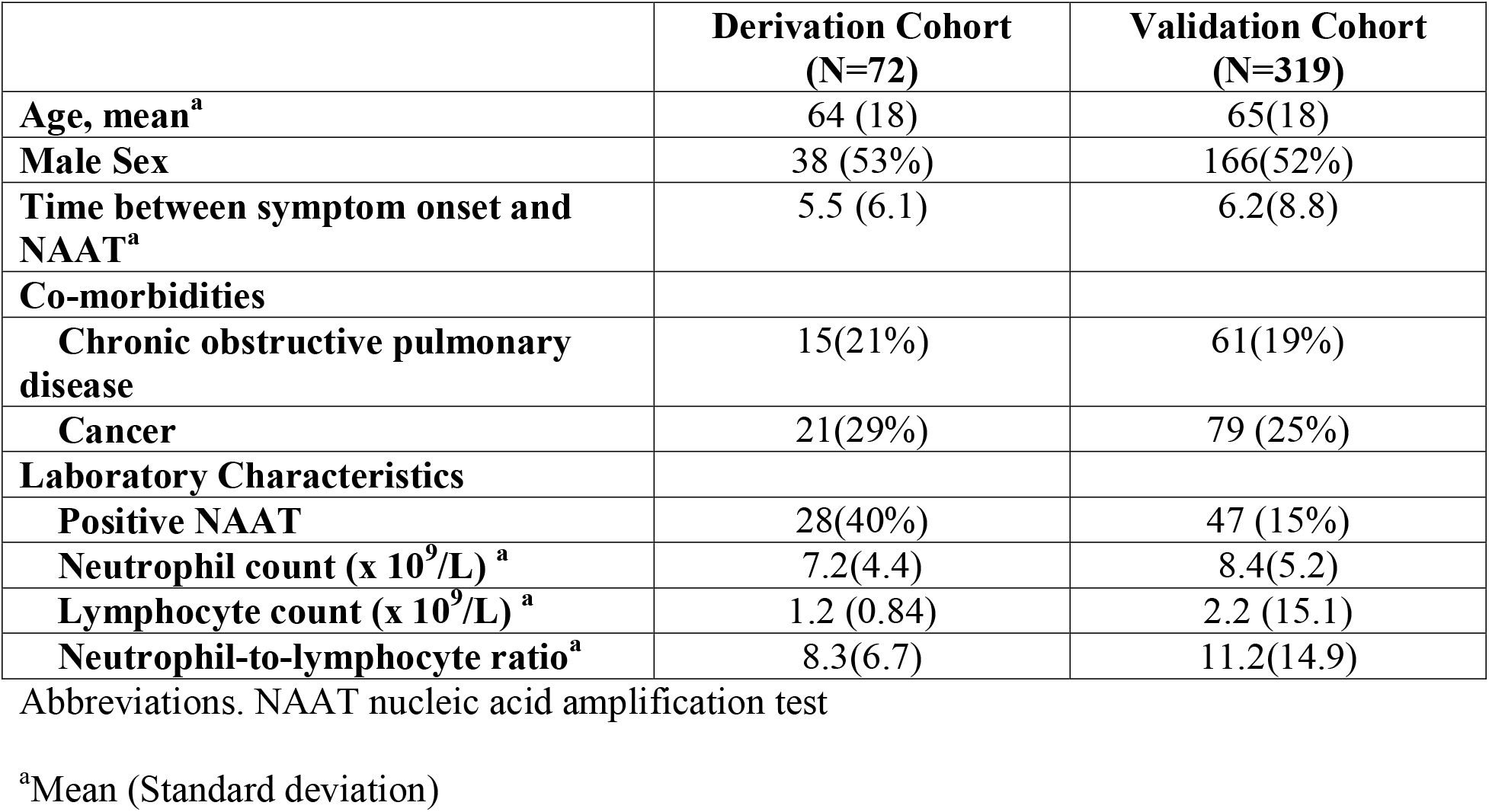
Population characteristics.

### 3.2. Performance of serology-based assays

In the derivation cohort serology testing was performed using Elecsys Anti-SARS-CoV-2 (N=57), EUROIMMUN Anti-SARS-CoV-2 ELISA IgA (N=59), EUROIMMUN Anti-SARS-CoV-2 ELISA IgG (N=72), DiaSorin’s LIAISON SARS-CoV-2 S1/S2 IgG (N=59), and Epitope Diagnostics Novel Coronavirus COVID-19 IgM (N=38). Diagnostic performance for the cut-off points as determined by ROC curve analysis and results of logistic regression analysis are shown in **Table 2**. All cut-off points identified were below the negative reference value for each reagent, except for the Euroimmun IgA assay. Of the included reagents, the Elecsys Anti-SARS-CoV-2 showed the best performance in primary and sensitivity analysis with the highest β-coefficient in the latter. The validation cohort was then tested with the Elecsys Anti-SARS-CoV-2 reagent. Using ROC curve analysis we estimated the diagnostic performance for 3 cut-off points: 0.1 (as estimated in the derivation cohort), 0.114 (estimated from the whole validation cohort), and 0.095 (estimated for the derivation cohort samples with a time differential between NAAT and serology within 48 hrs.). Results are shown in **Table** 3. At any given cut-off point the NPV was over 97%.

**Table 2.** Performance characteristics for serology assays

| **Reagent** | **Cut-off value** | **Sensitivity** | **Specificity** | **-LR** | **NPV<sup>a</sup>** | **Exp($\beta$)<sup>b</sup>** | **95% CI** | **Sig.** | **Exp($\beta$)<sup>b</sup>** | **95% CI** | **Sig.** |
| --- | --- | --- | --- | --- | --- | --- | --- | --- | --- | --- | --- |
| **Elecsys (N=57)** | 0.1 | 76.9 | 96.8 | 0.24 | 98.8 | 77.2 | 11.7-510.5 | <0.001 | 66 | 3.5-1254.6 | 0.005 |
| **Euroimmun IgA (N=59)** | 1.94 | 76.2 | 89.5 | 0.27 | 98.6 | 19.3 | 4.3-85.3 | <0.001 | 13.5 | 1.34-134 | 0.027 |
| **Euroimmun IgG (N=72)** | 0.445 | 67.9 | 90.9 | 0.35 | 98.2 | 22.4 | 5.7-87.3 | <0.001 | 17.5 | 2.3-134-3 | 0.006 |
| **Diasorin IgG (N=59)** | 5.7 | 75.0 | 97.7 | 0.26 | 98.7 | 126.5 | 11.9-1342.9 | <0.001 | NE | NE | NE |
| **Epitope IgM (N=38)** | 0.153 | 83.3 | 95.0 | 0.18 | 99.1 | 116.6 | 8.2-1661.6 | <0.001 | NE | NE | NE |

Abbreviations. -LR negative likelihood ratio; NPV negative predictive value; NE not estimable
<sup>a</sup>Values assume a 5% prevalence of positive tests.
<sup>b</sup>Adjusted for Time differential between NAAT and serology testing
<sup>b</sup>Subgroup analysis in samples with a time differential between NAAT and serology testing within 48 hrs.

**Table 3.** Performance characteristics of the Elecsys Anti-SARS-CoV-2 in the validation cohort

| Cut-off value | Sensitivity | Specificity | -LR | NPV<sup>a</sup> | Exp(b)<sup>b</sup> | 95% CI | Sig. |
| --- | --- | --- | --- | --- | --- | --- | --- |
| **Whole Cohort** |  |  |  |  |  |  |  |
| 0.1 | 91.5 | 89.3 | 0.1 | 99.5 | 90.7 | 26.0-316.2 | <0.001 |
| 0.114 | 87.2 | 95.6 | 0.13 | 99.3 | 118.7 | 40.8-345.6 | <0.001 |
| **Sensitivity analysis in samples with a time differential between NAAT and serology testing within 48 hrs.** |  |  |  |  |  |  |  |
| 0.095 | 75 | 86.18 | 0.29 | 98.5 (5%)<br>99.1 (3%)<br>99.7 (1%) | 21.5 | 2.0-228.2 | 0.011 |
| 0.1 | 50 | 93.4 | 0.54 | 97.3 (5%)<br>98.4(3%)<br>99.5(1%) | 14.2 | 1.1.8-111.7 | 0.012 |
| 0.114 | 66.7 | 96.05 | 0.35 | 98.2(5%)<br>98.9(3%)<br>99.7(1%) | 20.8 | 2.4-179.0 | 0.006 |

Abbreviations. -LR negative likelihood ratio; NPV negative predictive value; NE not estimable
<sup>a</sup>Values in brackets indicate disease prevalence assumptions. Default assumption is 5%.
<sup>b</sup>Adjusted for Time differential between NAAT and serology testing

## 4. Discussion

Despite advances in serology-based testing for SARS-CoV-2 infection, NAAT-based testing remains the gold standard but comes with disadvantages of availability, high cost and slower turn-around-time. These limitations are particularly important in resource-constrained settings. Over 18 months since the onset of the COVID-19 pandemic, as cases of COVID-19 continue to spread worldwide there remains a strong need for rapid, readily available diagnostic testing for SARS-CoV-2 infection. In this study, our findings suggest that by lowering the manufacturers’ recommended cut-off values for commercially available anti-SARS-CoV-2 antibodies serology we can accurately predict negative NAAT testing. In our population we would have had 77.4% (95% CI 72.5-81.7) of the samples categorized as highly likely to be negative by NAAT testing. Based on this data, we propose a ‘serology-first’ diagnostic algorithm that can be used to rule out SARS-CoV-2 infection in unvaccinated *symptomatic* patients presenting to the ED. This serology-based diagnostic algorithm offers a lower cost alternative to universal NAAT testing, in addition to rapid turnaround times to enable efficient triage, clinical management and infection control. Such an algorithm could be used to facilitate more judicious use of NAAT-based testing, contributing to more effective utilization of hospital resources, minimize healthcare worker exposure risk and nosocomial spread.

These findings could have significant implications for the diagnosis and management of SARS-CoV-2 infection in acute care settings. The high NPV of the serologic testing would not only reduce the need for NAAT testing but could allow early identification of the lower risk patients. Based on our observations, it could be reasonable to minimize isolation measures in serologically negative patients with a low probability of having a positive NAAT result such as those presenting with non-febrile illnesses. However, for others who have other indications for isolation such as respiratory tract symptoms with fever, isolation would still be required due to the risk of other respiratory pathogens.

The proposed approach has the additional advantage of using enzymatic methods that are standard for multiple applications in medicine and are widely available. In addition to a valuable tool for clinicians, an algorithm may find widespread application given the limited availability of NAAT-based testing especially in resource-constrained areas, in which NAAT testing may not be available, or the turn-around time for the results may be delayed by several days. Based on our observations, it could be reasonable to minimize isolation measures in patients with a low probability of having a positive NAAT result.

The current approach has some disadvantages, the most important of which are that it only applies to unvaccinated populations, and that its performance depends on the prevalence of positive SARS-CoV2 cases in the population in which it will be applied. As vaccination coverage increases, the performance of this method is yet to be determined but given the large disparities observed in many countries we believe that it still has a valuable place in clinical practice. Our study has some potential limitations. Our cohort was limited to patients who presented to the emergency department and were ill enough to require blood tests. It is unclear if the performance will be equivalent in a community setting such as an assessment center in which blood testing might have been deemed unnecessary. Another potential limitation is the fact that not all samples were obtained on the same day as the NAAT testing was performed. However, exhaustive sensitivity analyses confirmed robust results for this approach. Another consideration is that the derivation phase of this study showed that although potentially the use of different reagents might be useful in theory, inherent differences in the performance and characteristics of each test are likely going to affect the usefulness of our approach. The validated reagent detects total antibodies directed against the nucleocapsid of the SARS-CoV2 which are the ones that are elevated first during the immune response to the virus(7). That might explain to some extent the difference in the performance compared to other assays which detect antibodies against the spike protein. We hypothesize that given that our approach is only applicable to symptomatic patients, and that the mean duration of symptoms in this study was around 5 days, the early phase immune response mounting at the time of testing would not reach levels to meet the manufacturer’s diagnostic threshold, but by lowering the threshold the performance of the test can be modified. In other words, lowering the threshold of these tests will not help to identify SARS-CoV2 positive symptomatic patients, but can help to identify negative ones. This approach needs to be further explored using alternative automated or semi-automated platforms. Finally, our cohorts were assessed prior to the emergence of the delta wave in Canada. It has been reported that the viral load rises more rapidly and earlier in delta infections than in previous strains, although the time to symptom onset is less clear(8). Whether the serological results reported here in early infection will perform as well in patients infected with the delta strain will require further study. In summary, this study suggests that a SARS-CoV2 diagnostic strategy using the Elecsys Anti-SARS-CoV-2 serology test in the initial evaluation of symptomatic unvaccinated patients could be of clinical value to initially rule out an acute SARS-CoV2 infection and could potentially lead to appropriately tailored infection control measures or rational guidance for further testing with a potential reduction in costs and increased availability. The adequacy of this strategy with other reagents and in areas with a very high prevalence of positive cases needs further research.

## Data Availability

All data produced in the present study are available upon reasonable request to the authors

## Authors’ contributions

AL-L, BCY, ICY, MK, JD, VB and MS were involved in study design. JT, LL and BCY were involved in data extraction. BDH, JH, VB, and MK were involved in laboratory analysis. AL-L was involved in data analysis and manuscript draft. All authors provided revisions to the final manuscript.

## Acknowledgements

Authors wish to thank Husam Abdoh for helping with sample management.

